# Decreased cerebral blood flow in non-hospitalized adults who self-isolated due to COVID-19

**DOI:** 10.1101/2022.05.04.22274208

**Authors:** William S.H. Kim, Xiang Ji, Eugenie Roudaia, J. Jean Chen, Asaf Gilboa, Allison Sekuler, Fuqiang Gao, Zhongmin Lin, Aravinthan Jegatheesan, Mario Masellis, Maged Goubran, Jennifer S. Rabin, Benjamin Lam, Ivy Cheng, Robert Fowler, Chris Heyn, Sandra E. Black, Simon J. Graham, Bradley J. MacIntosh

**Author notes:** **Corresponding Author:** Bradley J. MacIntosh, Sunnybrook Research Institute, University of Toronto, 2075 Bayview Avenue, Room M6-180, M4N 3M5, Toronto, ON, Canada. indicates senior authorship (Simon J. Graham, Bradley J. MacIntosh).

## Abstract

The long-term consequences of coronavirus disease 2019 (COVID-19) on brain physiology and function are not yet well understood. From the recently described NeuroCOVID-19 study, we examined cerebral blood flow (CBF) in 50 participants recruited to one of two groups: 1) adults who previously self-isolated at home due to COVID-19 (n = 39; 116.5 ± 62.2 days since positive diagnosis), or 2) controls who experienced flu-like symptoms but had a negative COVID-19 diagnosis (n = 11). Participants underwent arterial spin labeling magnetic resonance imaging at 3 T to yield measures of CBF. Voxel-wise analyses of CBF were performed to assess for between-group differences, after controlling for age and sex. Relative to controls, the COVID-19 group exhibited decreased CBF in the thalamus, orbitofrontal cortex, and regions of the basal ganglia. Within the COVID-19 group, CBF differences in occipital and parietal regions were observed between those with (n = 11) and without (n = 28) self-reported on-going fatigue. These results suggest long-term changes in brain physiology in adults across the post-COVID-19 timeframe. Moreover, CBF may aid in understanding the heterogeneous symptoms of the post-COVID-19 condition. Future longitudinal studies are needed to further characterize the consequences of COVID-19 on the brain.

## 1. Introduction

Growing evidence suggests that the consequences of severe acute respiratory syndrome coronavirus 2 (SARS-CoV-2) infection extend beyond the respiratory system.^1,2^ As many as two thirds of individuals suffering from coronavirus disease 2019 (COVID-19) are reported to experience neurological and/or psychiatric symptoms during acute stages of infection.^3–5^ In some cases, symptoms have been reported to persist or even develop in the months following infection;^6^ this stage of COVID-19 has been referred to as the “post-COVID-19 condition” by the World Health Organization. Symptoms such as fatigue and so-called “brain fog” prevail in the post-COVID-19 timeframe;^2,7,8^ however, the long-term impact of COVID-19 on the brain is not well characterized. Efforts aimed at describing the post-COVID-19 condition as it relates to the brain are needed to mitigate pressure on strained healthcare systems worldwide.^9^

The effects of SARS-CoV-2 infection on the central nervous system are complex and likely involve multiple potential pathways. One theorized pathway is the nasal mucosal route of entry, whereby the virus may travel from the olfactory bulb to the primary olfactory cortex, which has direct connections to several brain regions including the thalamus, orbitofrontal cortex, and other midbrain regions;^10^ however, conclusive evidence of this pathway remains elusive. Another potential pathway may involve SARS-CoV-2 infiltrating cells expressing the angiotensin-converting enzyme (ACE-2) receptor, notably endothelial cells of the vasculature,^11^ and thus constituents of the neurovascular unit. For example, stroke and cerebrovascular disease secondary to COVID-19 have been reported.^12,13^ This notion of neurovascular involvement is further supported by a recent study demonstrating SARS-CoV-2 infection of tissue-cultured pericyte-like cells.^14^ In both cases, particularly the ACE-2 receptor pathway,^15^ SARS-CoV-2-induced neuroinflammation is likely to contribute to the post-COVID-19 condition.

Neuroimaging studies have shown that COVID-19 is associated with alterations to brain structure and/or punctate lesions (i.e., microbleeds, white matter hyperintensities), often in small samples of acutely infected individuals.^12,16–20^ In a unique study using pre- and post-infection data from a large UK Biobank sample, Douaud et al. observed longitudinal decreases in grey matter thickness, particularly in limbic regions, among adults who self-isolated or were hospitalized due to COVID-19.^21^ There are few cohort neuroimaging studies focusing on brain physiology in the post-COVID-19 timeframe;^1,22–28^ such imaging contrasts may be particularly relevant given the putative involvement of the vasculature in SARS-CoV-2 infection.^29,30^ Of the studies that exist, most involve adults who were hospitalized or in intensive care due to a more severe course of COVID-19.

As part of the Toronto-based NeuroCOVID-19 study,^31^ we conducted arterial spin labeling (ASL) magnetic resonance imaging (MRI) to probe cerebral blood flow (CBF), a measure of brain physiology and function, in non-hospitalized adults recovering from COVID-19. Our primary aim was to compare voxel-wise CBF (with and without partial volume correction) between adults who previously self-isolated at home due to COVID-19 and controls who experienced flu-like symptoms but tested negative for COVID-19. We hypothesized that the adults who previously self-isolated due to COVID-19 would exhibit altered CBF relative to controls, when assessed weeks/months beyond infection. Given the prevalence of fatigue as a symptom of the post-COVID-19 condition,^7,8,32–35^ we then performed an exploratory analysis of the association between self-reported fatigue and CBF among COVID-19 participants.

## 2. Materials and methods

### 2.1. Participants

Participants in the current study were recruited between May 2020 and September 2021 through the Department of Emergency Medicine at Sunnybrook Health Sciences Centre, physician referral, and community advertisements. Eligibility and consenting procedures were performed over phone or email. The Research Ethics Board at Sunnybrook Health Sciences Centre approved this study.

Inclusion criteria for this study included being between 20 and 75 years of age and having documented evidence of a positive or negative COVID-19 diagnosis, as determined by a provincially-approved facility through a nasopharyngeal and/or oropharyngeal swab and subsequent real-time reverse transcription polymerase chain reaction (PCR) test. Exclusion criteria for this study included previous diagnosis of dementia, an existing neurological disorder, previous traumatic brain injury, severe psychiatric illness, on-going unstable cardiovascular disease, or contraindications to MRI (e.g., ferromagnetic implants).

### 2.2. Study setting

Sunnybrook Health Sciences Centre is an academic tertiary level hospital. Prior to the pandemic, its emergency department received approximately 61,000 patients per year. Specialty services include trauma, interventional cardiology, stroke, oncology, neurosurgery, psychiatry, and high-risk obstetrics and gynecology. Its catchment area includes the Greater Toronto area and nearby regions (https://sunnybrook.ca/content/?page=care-programs).

### 2.3. Study design

The current study is an observational cohort neuroimaging study and is part of the NeuroCOVID-19 protocol, which has been previously described.^31^ We report on participants who were recruited to one of two groups: 1) adults who previously self-isolated at home due to COVID-19, or 2) controls who experienced flu-like symptoms but tested negative for COVID-19. Herein, we refer to the former as the COVID-19 group and the latter as the control group. The rationale for including this unique control group was that they may act as a better “baseline” against which the COVID-19 group could be compared (i.e., a group with non-specific flu-like symptoms who tested negative for COVID-19). Once non-infectious (i.e., following completion of a 14-day quarantine period and/or a negative PCR test), participants were invited for an on-site visit. Study staff and participants abided by the hospital’s infection prevention and control guidelines.

The primary outcome measure of the current study is ASL-derived CBF. Other outcome measures were assessed using: 1) a self-reported questionnaire of flu-like symptoms, 2) the Cognition and Emotion Batteries from the National Institutes of Health (NIH) Toolbox,^36,37^ and 3) the 40-odorant University of Pennsylvania Smell Identification Test (UPSIT, Sensonics International).^38^ The latter two assessments have been well-validated.^39–41^

The self-reported questionnaire of symptoms assessed whether participants were currently experiencing, had previously experienced, or had never experienced any flu-like symptoms including: fever, cough, sore throat, shortness of breath, fatigue, gastrointestinal symptoms, and/or smell/taste changes. Study staff ensured that symptoms were understood as being impairing to activities of daily living.

The Cognition Battery from the NIH Toolbox resulted in two age-corrected standard scores (mean = 100, standard deviation = 15) of fluid and crystallized cognition. The Emotion Battery resulted in three T-scores (mean = 50, standard deviation = 10) of negative affect, social satisfaction, and well-being. Note that a higher T-score for negative affect reflects more unpleasant moods and/or emotions. The interpretation of these scores has been previously described (https://nihtoolbox.force.com/s/article/nih-toolbox-scoring-and-interpretation-guide).

The UPSIT was administered as reports of olfactory dysfunction are a prevalent symptom of COVID-19.^7,8^ This assessment resulted in an UPSIT score (calculated as the number of odorants correctly identified) and a diagnosis of olfactory function (normosmia, mild hyposmia, moderate hyposmia, severe hyposmia to total anosmia). These diagnoses were determined as a function of UPSIT score and sex.

### 2.4. MRI acquisition

The MRI sequences used in this study consisted of T1-weighted and pseudo-continuous ASL acquired on a 3 T MRI system (Magnetom Prisma, Siemens Healthineers, Erlangen, Germany). T1-weighted images were acquired in three dimensions using an isotropic sagittal magnetization-prepared rapid gradient-echo sequence (TR/TE/TI = 2500/4.7/1100 ms, spatial resolution = 1 mm^3^, field-of-view = 256 mm, slices = 192, duration = 3:45 min:s). ASL images were acquired in three dimensions using an echo-planar turbo gradient-spin echo sequence with background suppression (TR/TE = 4100/36.8 ms, isotropic spatial resolution = 2.5 mm^3^, field-of-view = 240 mm, label duration = 1500 ms, post-label delay = 1800 ms, 7 control-label pairs, duration = 4:27 min:s).^42^ Proton-density ASL reference images were acquired with a TR of 4.1 s for CBF calibration.

### 2.5. MRI processing

MRI processing was performed using tools from the FMRIB Software Library (FSL, version 6.0.3).^43^ T1-weighted images were processed using *fsl_anat* with steps that included brain extraction, tissue segmentation, and non-linear registration to Montreal Neurological Institute (MNI) space.

ASL images were processed using *oxford_asl* with steps that included motion correction, spatial regularization,^44^ generation of control-tag difference images, voxel-wise calibration using the ASL reference image and assumed values from the literature,^45^ linear registration to structural space followed by non-linear registration to MNI space,^46^ and spatial smoothing with a Gaussian kernel of full-width at half maximum of 5 mm. The resulting CBF maps were then intensity-normalized (i.e., each CBF map was scaled to a global mean of 1) to account for between-participant global CBF differences.^47^ Two individuals (WSHK, BJM) visually inspected the CBF maps for quality control.

### 2.6. Statistical analysis

Demographic and clinical characteristics were compared between groups using independent samples t-tests for continuous data and chi-squared tests for categorical data. In cases when continuous data were non-normal (i.e., as assessed by the Shapiro-Wilk test), Mann-Whitney U-tests were used. In cases when categorical data had an expected value less than 5 (i.e., in a contingency table), Fisher’s exact tests were used. The threshold for statistical significance of demographic and clinical variables was set at 0.05.

For our primary aim, we performed between-group (i.e., COVID-19 vs. control) whole-brain voxel-wise analyses of CBF using two-tailed independent samples t-tests, controlling for age and sex. We used *3dFWHMx* and *3dClustSim* from the Analysis of Functional NeuroImages (AFNI, version 22.0.05) to estimate cluster-extent thresholds at a family-wise error rate of 0.05 with a cluster-forming threshold of 0.005. In addition, we performed one sensitivity analysis and one exploratory analysis in support of the primary aim.

#### Sensitivity analysis – Partial volume correction

We repeated the between-group comparison (i.e., COVID-19 vs. control) after including partial volume correction as an additional ASL processing step. The rationale for this sensitivity analysis is that ASL images were collected at a spatial resolution similar to the average thickness of the cortex, which may lead to biases in CBF estimation.^45^ Thus, due diligence was required to interpret CBF estimates and the resulting between-group differences. It is worth noting that currently available partial volume correction methods are inconsistent and may hinder interpretation; thus, it is recommended that partial volume correction be reported parallel to analyses using uncorrected CBF estimates.^45^ This additional processing step was implemented in *oxford_asl*.^48^

#### Exploratory analysis – Association between fatigue and CBF within the COVID-19 group

Given the prevalence of fatigue as a symptom of the post-COVID-19 condition,^7,8,32,33^ we examined whether COVID-19 participants who self-reported as experiencing on-going fatigue (n = 11) exhibited CBF differences compared to COVID-19 participants who previously reported fatigue that had resolved by the time of the assessment or did not experience fatigue at all (n = 28). Fatigue was determined using the self-reported questionnaire of symptoms.

## 3. Results

### 3.1. Demographic & clinical characteristics

At the time of analysis, a total of 50 participants (39 COVID-19, 11 controls) met eligibility criteria and had ASL and T1-weighted images available. Demographic and clinical characteristics are presented in Table 1.

**Table 1.**
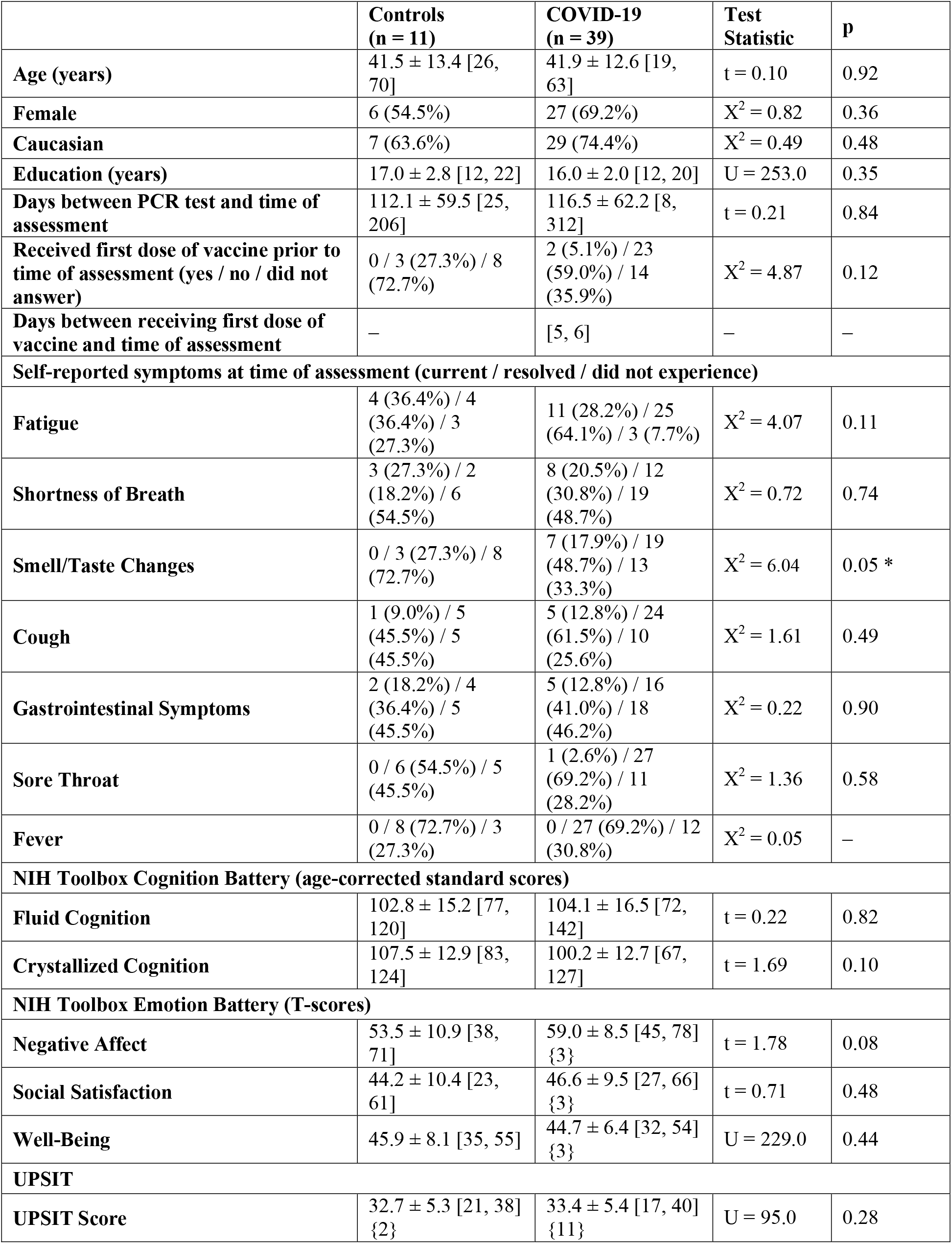
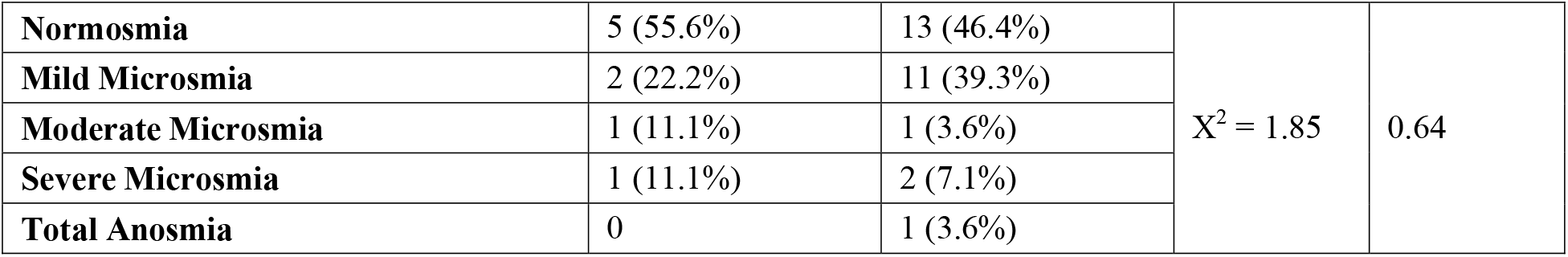
Demographic and clinical characteristics. Data are presented as mean ± standard deviation [minimum, maximum], or count (%). Between-group comparisons were performed using independent samples t-tests or Mann-Whitney U-tests for continuous data and chi-squared tests or Fisher’s exact tests for categorical data. Significant differences at p < 0.05 are indicated by an asterisk. Numbers in braces indicate participants with missing/faulty data. Abbreviations: PCR, polymerase chain reaction; NIH, National Institutes of Health; UPSIT, University of Pennsylvania Smell Identification Test.

Briefly, groups were well-matched for age and sex. COVID-19 participants were scanned 116.5 ± 62.6 [8, 312] days after receiving a positive diagnosis. Self-reported fatigue (COVID-19, 28.2%; control, 36.4%) and shortness of breath (COVID-19, 20.5%; control, 27.3%) were the most prevalent on-going symptoms across the cohort (Figure 1). Notably, 92.3% of COVID-19 participants and 72.7% of controls had experienced fatigue at some point between the PCR test and the time of the assessment. Significantly more COVID-19 participants had previously experienced or were currently experiencing smell/taste changes compared to controls (X^2^ = 6.04, p < 0.05). There were no between-group differences in fluid or crystallized cognition as assessed by the NIH Toolbox Cognition Battery, negative affect, social satisfaction, or well-being as assessed by the NIH Toolbox Emotion Battery (n.b., three COVID-19 participants did not complete the Emotion Battery), or UPSIT score (n.b., 11 COVID-19 participants and two controls had missing/faulty UPSIT data).

**Figure 1.**
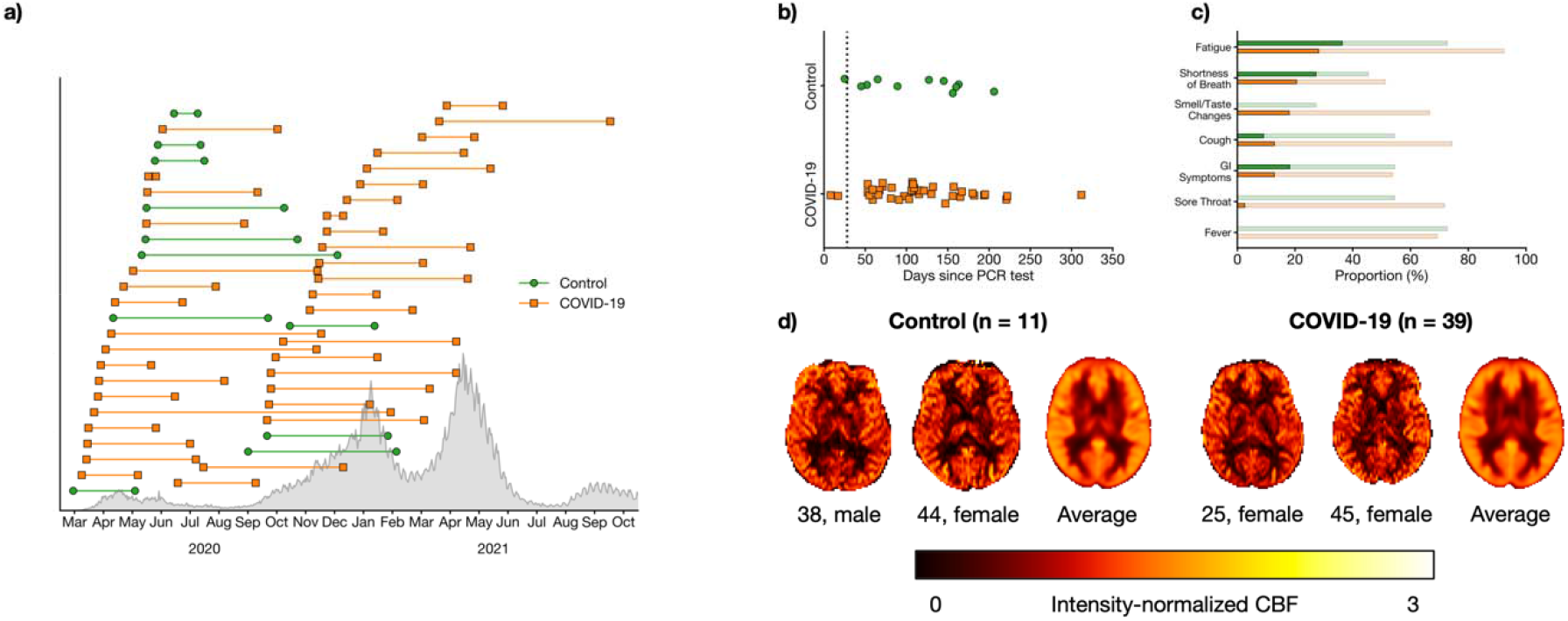
a) Timing of PCR test (left marker) and assessment (right marker) for COVID-19 (orange squares) participants and controls (green circle). Confirmed cases in Ontario are shown in grey. b) Number of days between PCR test and assessment. The black dotted line indicates 28 days, an established threshold beyond which symptoms can be considered part of the post-COVID-19 condition. c) Proportion of participants who self-reported flu-like symptoms. Faint bars indicate participants whose symptoms had resolved by the time of the assessment while dark bars indicate participants with on-going symptoms. d) Representative and group-averaged CBF maps from both groups.

### 3.2. Differences in CBF between COVID-19 and control groups

Relative to controls, the COVID-19 group exhibited significantly decreased CBF in a large cluster of voxels encompassing the thalamus, orbitofrontal cortex, and regions of the basal ganglia, including the caudate, nucleus accumbens, putamen, and pallidum (Figure 2 and Table 2). There were no clusters in which the COVID-19 group had significantly increased CBF relative to controls.

**Figure 2.**
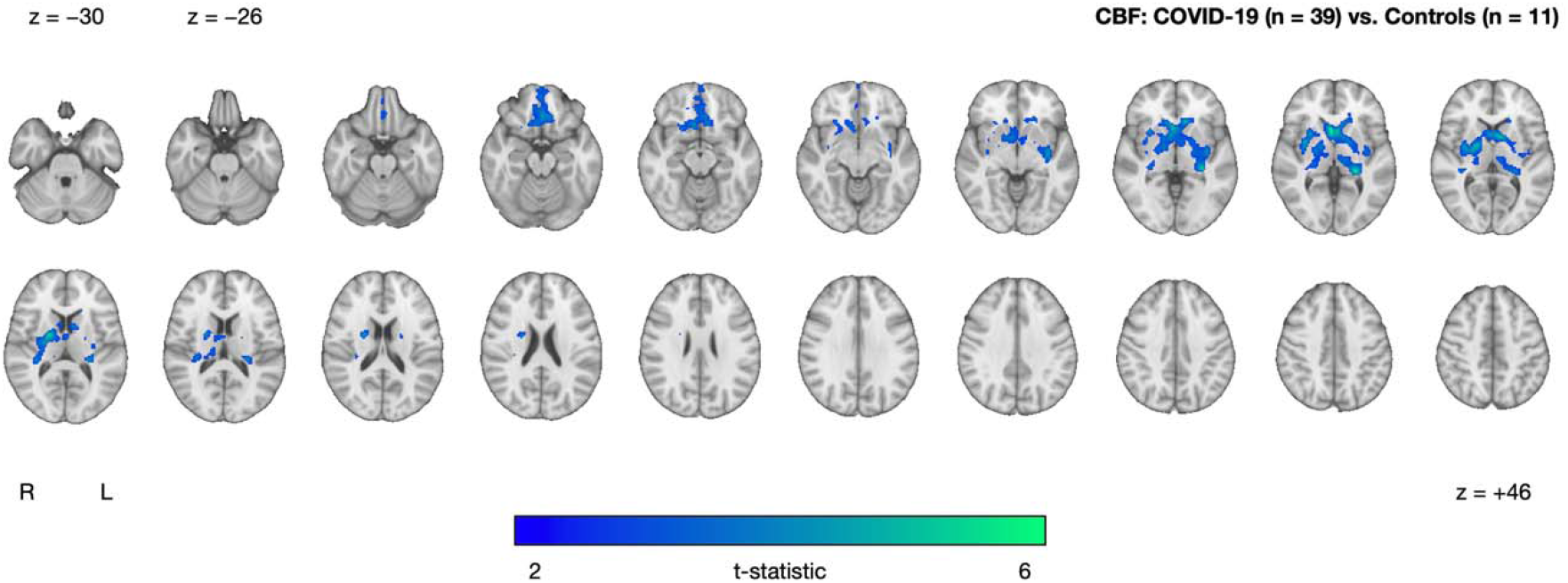
Cluster exhibiting significantly decreased CBF in the COVID-19 group (n = 39) relative to controls (n = 11), after controlling for age and sex. No clusters were found where the COVID-19 group had higher CBF compared to controls. Statistical maps are presented in radiological convention. Montreal Neurological Institute coordinates are denoted by z-values. Abbreviations: R, right; L, left.

**Table 2.** Summary of voxel-wise analyses of CBF. The primary (top row) and secondary analyses (bottom rows) show results of the two-tailed independent samples t-tests that were used to test for between-(sub)group differences, controlling for age and sex. Coordinates indicate location of peak t-statistic.

| Comparison | Direction | Size | t-statistic | x | y | z | Description |
| --- | --- | --- | --- | --- | --- | --- | --- |
| COVID-19 (n = 39) vs. Controls (n = 11) | COVID-19 < Controls | 4,431 | 5.94 | 4 | 14 | 2 | Thalamus, Orbitofrontal Cortex, Caudate, Nucleus Accumbens, Putamen, Pallidum |
| *Sensitivity Analysis – Partial Volume Correction* |  |  |  |  |  |  |  |
| COVID-19 (n = 39) vs. Controls (n = 11) with partial volume correction | COVID-19 < Controls | 2,251 | 4.83 | −4 | 8 | 6 | Thalamus, Orbitofrontal Cortex, Caudate, Nucleus Accumbens, Putamen, Pallidum |
| *Exploratory Analysis – Effects of Fatigue on CBF within the COVID-19 group* |  |  |  |  |  |  |  |
| COVID-19 with fatigue (n = 11) vs. COVID-19 without fatigue (n = 28) | COVID-19 with fatigue > COVID-19 without fatigue | 464 | 4.40 | 32 | −60 | 50 | Superior Lateral Occipital Cortex, Angular Gyrus, Superior Parietal Lobule, Supramarginal Gyrus |
|  | COVID-19 with fatigue < COVID-19 without fatigue | 758 | 4.75 | 10 | −66 | −4 | Lingual Gyrus, Occipital Fusiform Gyrus, Intracalcarine Cortex, Precuneous Cortex |

### 3.3. Sensitivity analysis – Differences in CBF between COVID-19 and control groups with partial volume correction

Our sensitivity analysis with partial volume correction resulted in a similar cluster of smaller extent compared to the primary analysis (Figure 3 and Table 2). Again, there were no clusters in which the COVID-19 group had significantly increased CBF relative to controls.

**Figure 3.**
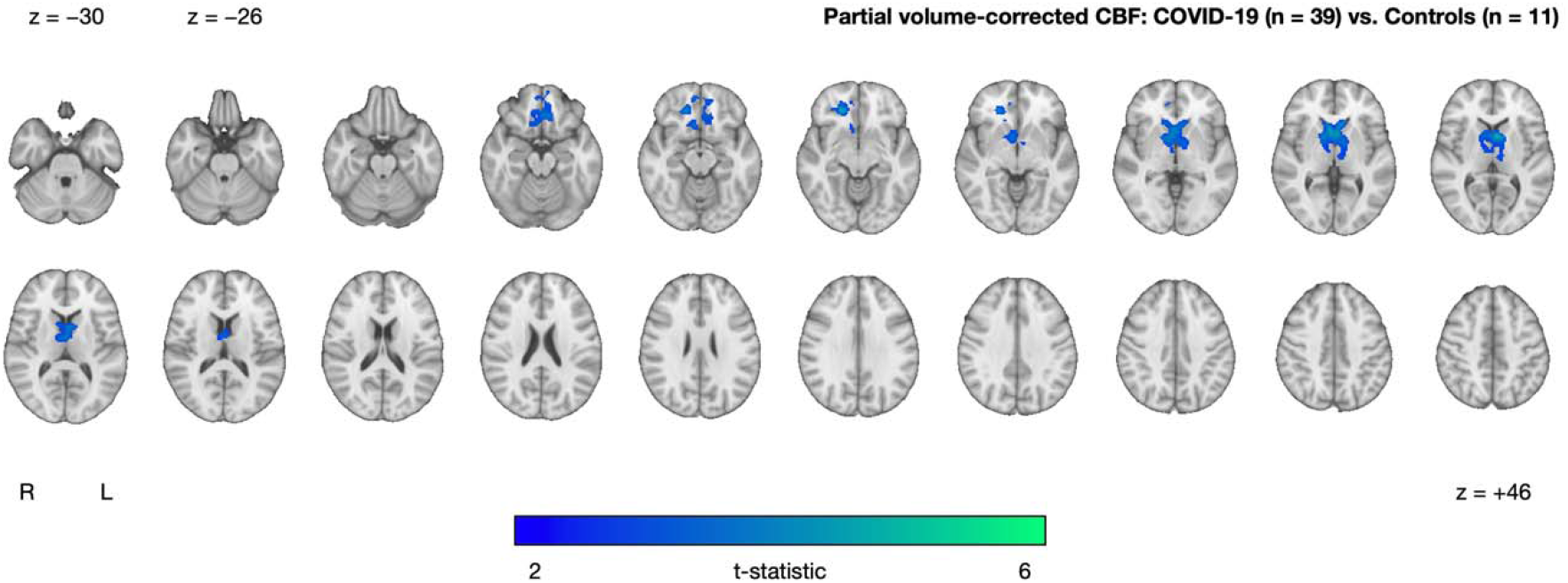
Cluster exhibiting significantly decreased partial volume-corrected CBF in the COVID-19 group (n = 39) relative to controls (n = 11), after adjusting for age and sex. No clusters were found where the COVID-19 group had higher CBF compared to controls. Statistical maps are presented in radiological convention. Montreal Neurological Institute coordinates are denoted by z-values. Abbreviations: R, right; L, left.

### 3.4. Exploratory analysis – Association between fatigue and CBF within the COVID-19 group

Within the COVID-19 group, we observed between-subgroup CBF differences between those with and without on-going fatigue. On-going fatigue was characterized by a cluster of increased CBF in superior occipital and parietal regions (superior lateral occipital cortex, angular gyrus, superior parietal lobule, supramarginal gyrus) and a cluster of decreased CBF in inferior occipital regions (lingual gyrus, occipital fusiform gyrus, intracalcarine cortex, precuneous cortex) (Figure 4 and Table 2).

**Figure 4.**
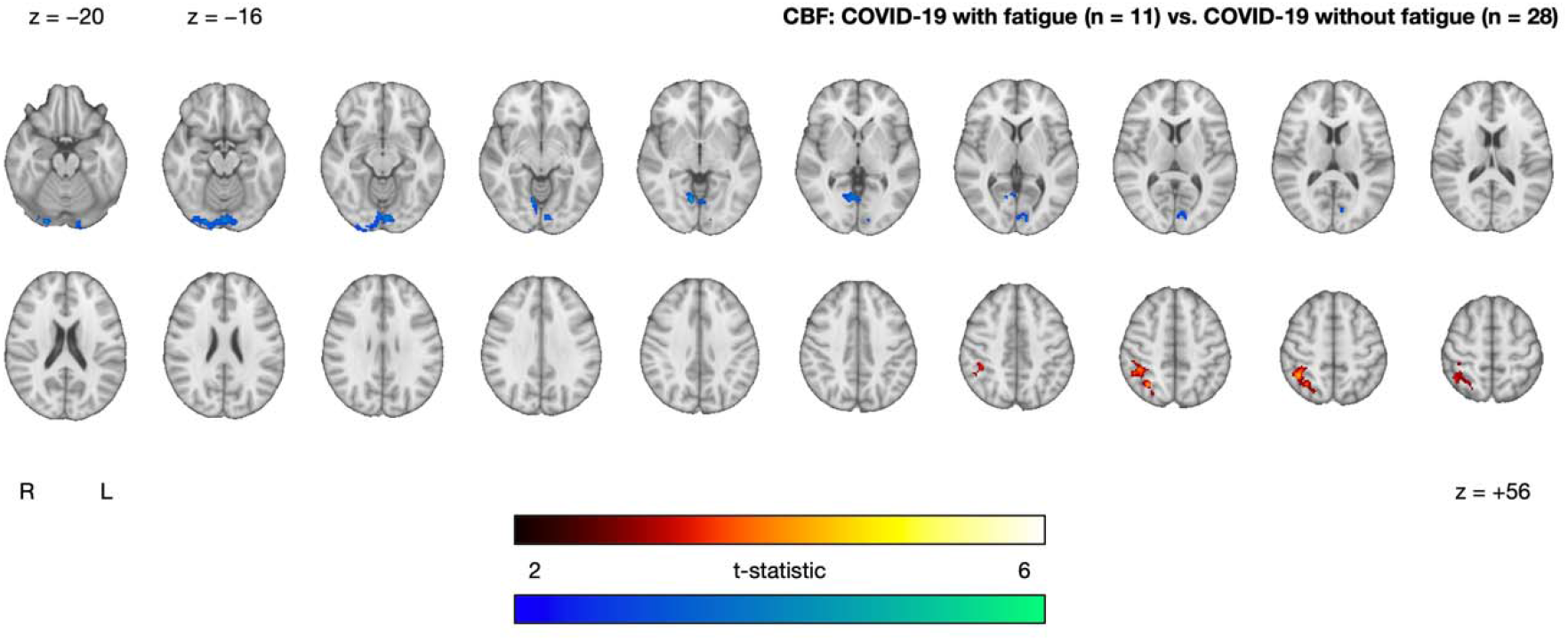
Clusters exhibiting significantly increased (red-yellow) and decreased (blue-green) CBF in the COVID-19 with fatigue group (n = 11) relative to the COVID-19 without fatigue group (n = 28), after controlling for age and sex. Statistical maps are presented in radiological convention. Montreal Neurological Institute coordinates are denoted by z-values. Abbreviations: R, right; L, left.

## 4. Discussion

In this study, we investigated whether adults who previously self-isolated at home due to COVID-19 would exhibit alterations in CBF when compared against controls who experienced flu-like symptoms but tested negative for COVID-19. COVID-19 participants exhibited significantly decreased CBF in the thalamus, orbitofrontal cortex, and regions of the basal ganglia compared to controls. We further examined the effect of fatigue within the COVID-19 group, which revealed between-subgroup CBF differences in occipital and parietal regions. These results provide support for long-term changes in brain physiology in adults across the post-COVID-19 timeframe.

Although COVID-19 is primarily a respiratory illness, the cerebrovasculature is also susceptible to damage as endothelial cells and pericytes are prone to viral invasion.^14,29^ The brain’s vasculature interfaces with the complex neurovascular unit, for instance, in the regulation of CBF.^49^ Furthermore, the location of potential brain involvement in relation to SARS-CoV-2 is likely to vary regionally, with some evidence to suggest that relative to the rest of the brain, ACE-2 receptor expression is highest in the thalamus, the paraventricular nuclei of the thalamus, and more generally in regions proximal to the ventricles.^50^ Notably, we found significantly decreased CBF in the anterior thalamus, which contains the paraventricular nuclei of the thalamus, a key region of the brain’s anxiety network.^51^ Moreover, decreased thalamic glucose metabolism, as measured by positron emission tomography (PET), has been observed at both acute and chronic stages of recovery from COVID-19.^26,27,52^

Decreased CBF was also detected in regions of the basal ganglia, including the caudate, nucleus accumbens, putamen, and pallidum. In particular, the caudate has been reported in a longitudinal PET study that observed decreased glucose metabolism in seven adults recovering from COVID-19, up to 6 months post-infection.^27^ Multivariate methods have also revealed that glucose metabolism within the caudate is a distinguishing feature between COVID-19 patients and controls.^24^ We also observed decreased CBF within the orbitofrontal cortex, a region widely reported as being associated with SARS-CoV-2 infection.^12,21,53–56^ Together with the thalamus and regions of the basal ganglia, the orbitofrontal cortex is a key region of the cortico-basal ganglia-thalamic loop, a circuit involved in complex behaviours including affect regulation and reward-based decision-making,^57^ as well as in relation to neurological and psychiatric disorders.^58,59^ Moreover, the orbitofrontal cortex also plays an important role in olfaction and is often referred to as the secondary olfactory cortex.^60^ The results of the current study align with previous PET studies that find decreased glucose metabolism within the orbitofrontal cortex, and more generally within the frontal lobe. In an early case report of one healthy 27-year-old with COVID-19 experiencing persistent anosmia, Karimi-Galougahi et al. reported decreased glucose metabolism in the left orbitofrontal cortex.^55^ Hosp et al. reported frontoparietal hypometabolism in 10 out of 15 adults with subacute COVID-19.^24^ Guedj et al. reported frontal hypometabolism in 35 adults that were 3 weeks beyond infection, and that significant clusters were correlated with higher occurrence of symptoms, such as anosmia.^26^ Finally, Kas et al. reported a consistent pattern of orbitofrontal, dorsolateral, and mesiofrontal hypometabolism in seven adults with acute COVID-19-related encephalopathy, despite heterogenous symptomatology, and posited that COVID-19 is related to frontal lobe impairment.^27^ Notably, the results from the latter study persisted until 6 months following infection. Altogether, the result of decreased CBF within the orbitofrontal cortex, along with the thalamus and regions of the basal ganglia, may reflect COVID-19-related disturbances to brain networks, olfactory function, and emotional/cognitive concerns. Future studies extending these potentially brain network-related results through investigations of functional and structural connectivity are warranted.

It is important to note that participants in the current study were recruited over the course of several pandemic waves in Ontario, each being associated with a different distribution of variants of concern (Figure 1). Thus, it is probable that COVID-19 participants were infected with different strains of SARS-CoV-2, likely spanning from the Alpha variant to the Delta variant. We further note that these participants were recruited prior to the emergence of the Omicron variant which, despite its high transmissibility, is believed to be less severe than previous strains.^61,62^

Our comparison of COVID-19 participants with and without fatigue resulted in between-subgroup CBF differences, primarily in occipital and parietal regions of the brain. There have been efforts to characterize COVID-19 based on symptoms, with the hope of predicting severity and likelihood of the post-COVID-19 condition.^32,33^ Others have observed fatigue-related differences in brain structure and function in those recovering from COVID-19,^35^ such as functional connectivity alterations in parietal regions.^34^ Interestingly, the post-COVID-19 condition shares many common features with chronic fatigue syndrome (i.e., myalgic encephalomyelitis), a disorder that can be triggered by viral infection,^63^ and that is characterized by decreased CBF, such as within the lingual gyrus.^64,65^ Therefore, these fatigue-related CBF differences amongst COVID-19 participants could help guide therapeutic efforts in treating fatigue as a symptom of the post-COVID-19 condition. We note that while brain-behaviour investigations in the context of COVID-19 are important in understanding symptoms, this fatigue-related analysis is a “scratch of the surface”. Higher-order multivariate analyses (e.g., principal component analysis) with larger sample sizes will be better poised to answer such questions.

These results need to be interpreted in the context of several limitations. First, although well-matched, the sample sizes of the two groups were modest and unequal; furthermore, a power analysis was not performed. To our knowledge, the current study benefits from the largest ASL dataset focusing on non-hospitalized adults in the post-COVID-19 timeframe. Moreover, recruitment for the NeuroCOVID-19 study is on-going and will address these issues in future studies. Second, our recruitment may be confounded by selection bias. For example, the current study’s cohort was comprised of 66% female and 72% Caucasian participants. We further note that participants needed internet access to be screened for eligibility. Third, our control group exhibited flu-like symptoms of unknown origin. The recruitment of this unique control group is a relatively novel aspect of this study, as these participants are a de-novo sample of adults that experienced non-specific flu-like symptoms during the pandemic. Fourth, ASL images were acquired at a spatial resolution comparable to the average thickness of the cortex, which may be susceptible to partial volume error.^45^ To address this, we included partial volume correction as an additional ASL processing step in a sensitivity analysis, which did not drastically change the results. Fifth, our fatigue-related exploratory analysis relied on self-reported symptoms. Study staff ensured that on-going fatigue was understood as being impairing to activities of daily living. Finally, the data used in this study are cross-sectional and lack a pre-infection assessment.^21^ Further investigation into longitudinal changes of these participants will be performed as part of the NeuroCOVID-19 study. It may also be feasible to access pre-pandemic repository data from age- and sex-matched individuals.

In conclusion, we observed decreased CBF in those recovering from COVID-19 relative to controls. These decreases were present months after acute infection and were localized to regions that have previously been highlighted as related to SARS-CoV-2 infection. We also observed CBF differences in relation to fatigue within the COVID-19 group, suggesting that CBF may aid in parsing the heterogeneous symptoms associated with the post-COVID-19 condition. In all, these results suggest that the post-COVID-19 condition may be associated with long-term effects on brain physiology and function. Future studies that replicate and further characterize such effects are warranted.

## Data Availability

All data produced in the present study are available upon reasonable request to the authors.

## Acknowledgments

The authors wish to thank all study participants and staff (Ellen Cohen, Garry Detzler, Ruby Endre, Haddas Grosbein, Masud Hussain, Devin Sodums) for their time and contributions to this study. We thank Dr. Danny J.J. Wang from the University of Southern California for providing the 3D pCASL sequence.

## Authors’ contributions

Study design: WSHK, XJ, ER, JJC, AG, AS, FG, ZL, AJ, MM, MG, JR, BL, IC, RF, CH, SEB,SJG, BJM. Data collection: XJ, ER, ZL, AJ, SJG, BJM. Data analysis and interpretation: WSHK, XJ, ER, JJC, AG, AS, FG, ZL, AJ, MM, MG, JR, BL, IC, RF, CH, SEB, SJG, BJM. Manuscript writing: WSHK, XJ, ER, JJC, ZL, SJG, BJM. All authors revised and approved the final version of this manuscript.

